# Prevalence of Diabetes and Prediabetes in Peru: A Systematic Review and Meta-Analysis

**DOI:** 10.1101/2024.05.21.24307705

**Authors:** Víctor Juan Vera-Ponce, Fiorella E. Zuzunaga-Montoya, Luisa Erika Milagros Vásquez-Romero, Joan A. Loayza-Castro, Enrique Vigil-Ventura, Willy Ramos

## Abstract

**Introduction:** Diabetes mellitus (DM) and prediabetes represent significant public health challenges in Peru, with prevalence rates showing considerable variation in recent studies.

**Objective:** Conduct a systematic review and meta-analysis to determine the updated prevalence of diabetes and prediabetes in Peru.

**Methodology:** An exhaustive analysis of studies published in PubMed, Web of Science, SCOPUS, EMBASE, Scielo, and LILACS databases was conducted. Both national and subnational studies reporting prevalence rates of diabetes and prediabetes using different diagnostic methods and representative sampling techniques were included. Data were synthesized through meta-analysis, evaluating heterogeneity and trends over time and across regions.

**Results:** Eight studies provided data on the prevalence of DM, while six studies reported on prediabetes. The meta-analysis calculated an overall prevalence of DM at 5.90% (95% CI: 3.98% - 8.17%) and prediabetes at 13.02% (95% CI: 6.31% - 21.68%). Both conditions showed increasing trends over time, with recent studies indicating prevalence rates as high as 10%. However, there was substantial variability in prediabetes rates, ranging from 3.62% to 24.5%.

**Conclusions:** The findings underscore the need for standardization in diagnostic methods and the implementation of more extensive and representative national studies. To mitigate the impact of these chronic conditions, public health policies should focus on early detection and prevention.

## INTRODUCTION

Diabetes mellitus (DM) is a chronic disease that has seen a significant rise in prevalence globally over the past few decades, becoming a major public health concern ^(1)^. Notably, the prevalence of DM in developing countries has been growing faster than in developed nations ^(2)^. Peru is no exception in Latin America, with a steady increase in DM prevalence in recent years ^(3)^.

DM profoundly impacts individual health and places a substantial economic burden on health systems and national economies ^(4)^. Complications from DM, such as cardiovascular disease, kidney failure, and blindness, can severely decrease quality of life and increase mortality ^(5)^. Additionally, managing DM and its complications leads to significant healthcare costs, loss of productivity, and indirect costs related to disability and premature death ^(6)^. Therefore, effective prevention and control of DM are essential to improving public health and ensuring the sustainability of health systems in Peru.

Despite the rising prevalence of DM in Peru, accurately understanding its magnitude remains challenging. This difficulty is partly due to the lack of large-scale epidemiological studies and the variability in the quality of reported data. Recent research has shown significant disparities in prevalence figures, ranging from as low as 3%, as reported by the Nutritional Biochemical Indicators Survey (ENINBSC, acronym in Spanish) ^(7)^ or the study by Miranda JJ ^(8)^, to as high as 10% in the Nutritional and Food Surveillance by Life Stages (VIANEV, acronym in Spanish) study ^(9)^. This variability underscores significant inconsistencies in the estimates and highlights the uncertainty about the real impact of DM on the Peruvian population.

Although Carrillo and Bernabé conducted a review in 2019 ^(10)^, their study did not perform an exhaustive meta-analysis, excluded key data such as the VIANEV study, and did not include prediabetes. Therefore, the present study aims to conduct an updated systematic review with a meta-analysis of the prevalence of DM and prediabetes in Peru.

## METHODS

### Design

A systematic review (SR) with meta-analysis and pooled analysis of national cross-sectional studies was conducted, following the guidelines of the PRISMA statement (Preferred Reporting Items for Systematic Reviews and Meta-Analyses) ^(11)^, considering that this was an SR of prevalence studies.

### Additional Data Sources and Justification

In addition to published studies, technical reports and national databases were included for a more comprehensive evaluation. When reports and manuscripts were unavailable, data were extracted from national databases. In these cases, the original study that generated the database was cited to provide complete methodological context and ensure transparency in the data source.

### Search Strategy

From January 1 to March 3, 2024, a strategic search was conducted in six academic databases: Scopus, Web of Science, Embase, PubMed, LILACS, and Scielo. The key terms used for the search across all sources were “prevalence,” “diabetes,” “prediabetes,” and “Peru.” The detailed search strategy for each database and source of technical reports and national databases is provided in Supplementary Appendix 1.

### Selection Criteria

Studies were eligible for inclusion if they: 1) Were observational studies published as full articles or technical reports in the absence of full articles; 2) Included adult participants of both sexes; 3) Evaluated the prevalence of diabetes and prediabetes in Peru, with at least one blood marker, such as fasting glucose (FG), oral glucose tolerance test (OGTT), or glycated hemoglobin, as recommended by the American Diabetes Association ^(12)^ or the World Health Organization ^(13)^; 4) Used probabilistic sampling; and 5) Provided sufficient data to calculate prevalence or directly reported prevalence. Narrative reviews, editorials, preclinical studies, and studies not pertinent to the objective of this study were excluded.

### Study Selection

The Rayyan software (https://rayyan.qcri.org) was used to manage and store articles identified from each database. Two independent reviewers (FEZM and LEMVR) conducted an initial review of the titles and abstracts of manuscripts, technical reports, and national databases. If both reviewers agreed that a source met the inclusion criteria, it was selected for a more detailed review. In cases of disagreement, a third reviewer (WR) acted as an arbitrator.

Subsequently, a thorough full-text review of all preselected articles, reports, and databases was conducted. The decision to include or exclude each source was recorded in an Excel spreadsheet. This process was also performed by three reviewers, and any discrepancies were resolved by a fourth reviewer. For databases, original studies were consulted to understand the methodology and ensure that the data were appropriate for inclusion.

### Data Extraction and Qualitative Analysis

For each selected source, meticulous data extraction was performed using a Microsoft Excel 2016 template designed for this study. Extracted data included author, year of publication, study design, sample size, prevalence of diabetes and prediabetes, and blood markers used (such as fasting glucose, random glucose, glucose tolerance test, or glycated hemoglobin). Similar data were extracted for technical reports and national databases, and original studies were consulted to understand the methodology and ensure the quality of the extracted data. Additionally, measures of variability (such as the 95% confidence interval) were recorded when available.

### Risk of Bias Assessment

Two of our researchers independently assessed the risk of bias for all included studies using the Joanna Briggs Institute Checklist for Prevalence Studies ^(14)^. This tool was developed to increase consistency in systematic reviews of prevalence data and is recommended as the most appropriate for such studies ^(15)^.

Bias in studies was assessed using nine specific criteria: 1) Adequacy of the sampling frame to address the target population; 2) Appropriate selection of participants; 3) Appropriateness of the sample size; 4) Detailed description of the subjects and study context; 5) Adequacy of data analysis covering the identified sample; 6) Use of valid methods to diagnose the condition; 7) Standard and reliable measurement of the condition in all participants; 8) Adequacy of the statistical methods employed; 9) Proper handling of the response rate or its impact if low.

A score of 1 to 9 was considered based on whether the response was correct to evaluate the level of bias. Studies scoring 4 points or less were considered to have a high risk of bias. Those scoring between 5 and 6 points were classified as having a medium risk of bias, and studies scoring 7 points or more were categorized as having a low risk of bias.

### Quantitative Analysis

In this study, several statistical analyses were applied to evaluate the prevalence of diabetes and prediabetes in Peru using STATA version 18 software. First, a global meta-analysis of the prevalence of both conditions was conducted, integrating data from all selected studies to provide a consolidated estimate. This approach allowed for an overall view of the country’s situation and established a benchmark for more detailed comparisons.

Subsequently, sensitivity analyses were conducted to explore variations in prevalence according to different subgroups and periods. These analyses were structured around three main criteria: time, with a 10-year threshold before and after; geographic scope, differentiating between national and subnational studies; and sex, evaluating prevalence separately in male and female populations.

Additionally, heterogeneity was quantified using the I² statistic, which measures the percentage of total variation across studies due to heterogeneity rather than chance. 95% confidence intervals were calculated for all prevalence estimates.

## Results

### Eligible Studies

A total of 1108 publications were found. After removing duplicates (760), 348 manuscripts were analyzed based on title and abstract. After excluding 322 studies, 26 full-text articles were retrieved. Finally, after applying the selection criteria, 8 articles (3,7–9,16–19) were selected (see Figure 1).

**Figure 1.**
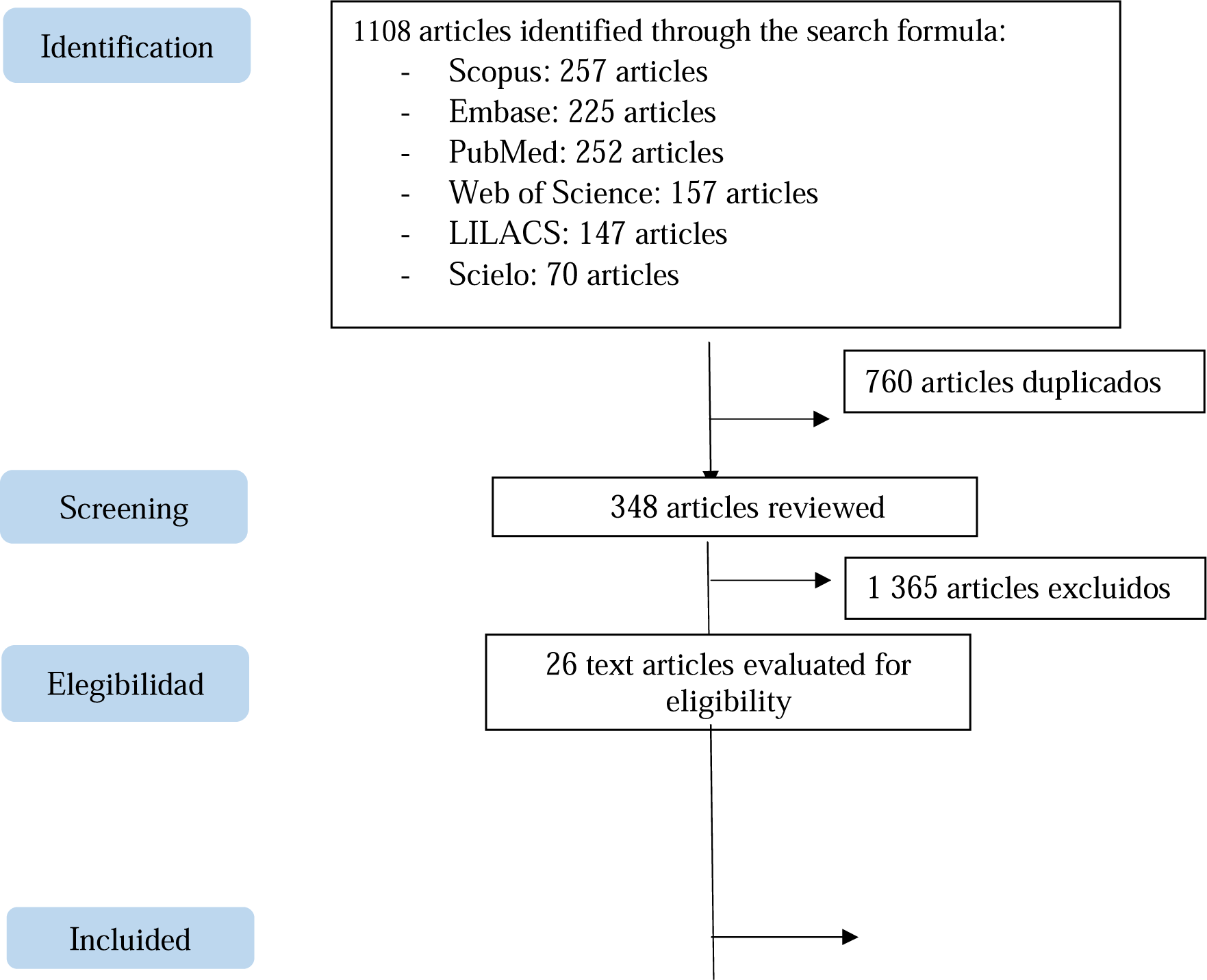
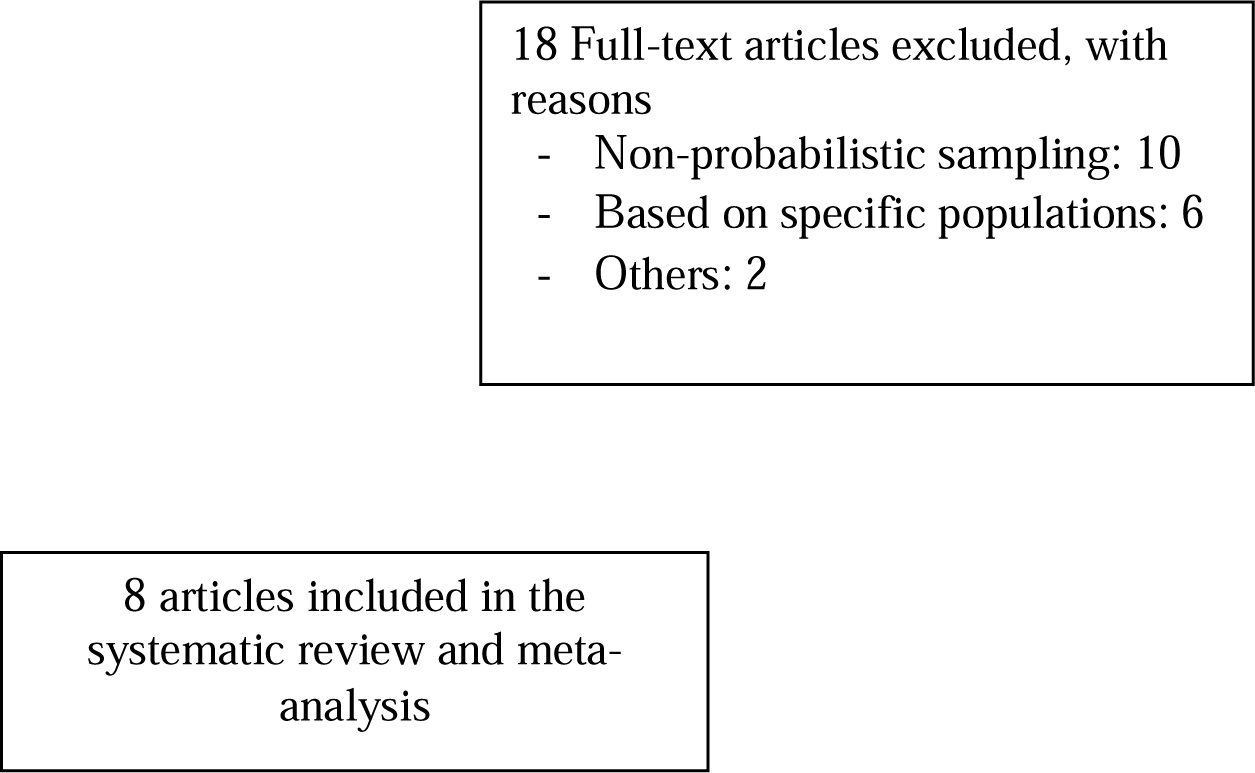
Flow Diagram of Study Selection.

### Characteristics of the Studies

The studies analyzed span a period from 2004 to 2018. Variations in sample size were observed, ranging from 989 participants in the study by Miranda JJ ^(8)^ to 3,238 in the study by Bernabé-Ortiz (2015) ^(16)^. A total of two national-level studies were identified, the ENINBSC^(7)^ and the VIANEV report ^(9)^, along with seven subnational studies, primarily focusing on specific regions such as Lima, Ayacucho, and Tumbes. Eight studies evaluated the prevalence of DM, while six evaluated prediabetes.

The studies showed considerable variation in terms of prevalence. Regarding DM, the VIANEV report recorded the highest prevalence at 10.7%, whereas the Miranda JJ ^(8)^ study reported the lowest at 3%. For prediabetes, the VIANEV report indicated a prevalence of 24.5%, while the ENINBSC report showed only 3.62%.

Regarding diagnostic methods, all studies used fasting glucose as the diagnostic criterion for DM, in addition to self-reporting. Bernabé-Ortiz’s (2018) ^(17)^ study also included the OGTT. All studies considered fasting glucose levels between 100 and 125 mg/dl for prediabetes, except for the VIANEV report, which defined prediabetes as fasting glucose levels above 110 mg/dl up to 125 mg/dl.

### Risk of Bias Assessment

The studies evaluated in this systematic review demonstrated high adherence to established criteria for minimizing the risk of bias, employing appropriate sampling frameworks, and effective recruitment methods. Each study ensured an adequate sample size and provided detailed descriptions of the subjects and study settings, which is essential for ensuring the validity and reliability of the results. Additionally, the methods used for identifying the health condition were consistently valid and reliable, and the data analysis was conducted with sufficient coverage of the identified sample, further strengthening the credibility of the reported prevalence estimates.

However, one of the studies’ most critical and variable aspects was handling response rates. Although not all studies provided explicit details on how potential low response rates were managed, those that did implemented appropriate measures to mitigate the impact on the results. This careful approach to managing response rates is crucial for maintaining a low risk of bias and ensuring that the findings are representative of the general population.

### Table 2. Meta-analysis of DM Prevalence

The pooled prevalence for DM was 5.90% (95% CI: 3.98%—8.17%). However, heterogeneity was high (I² = 96.97%) (Figure 2).

**Figure 2.**
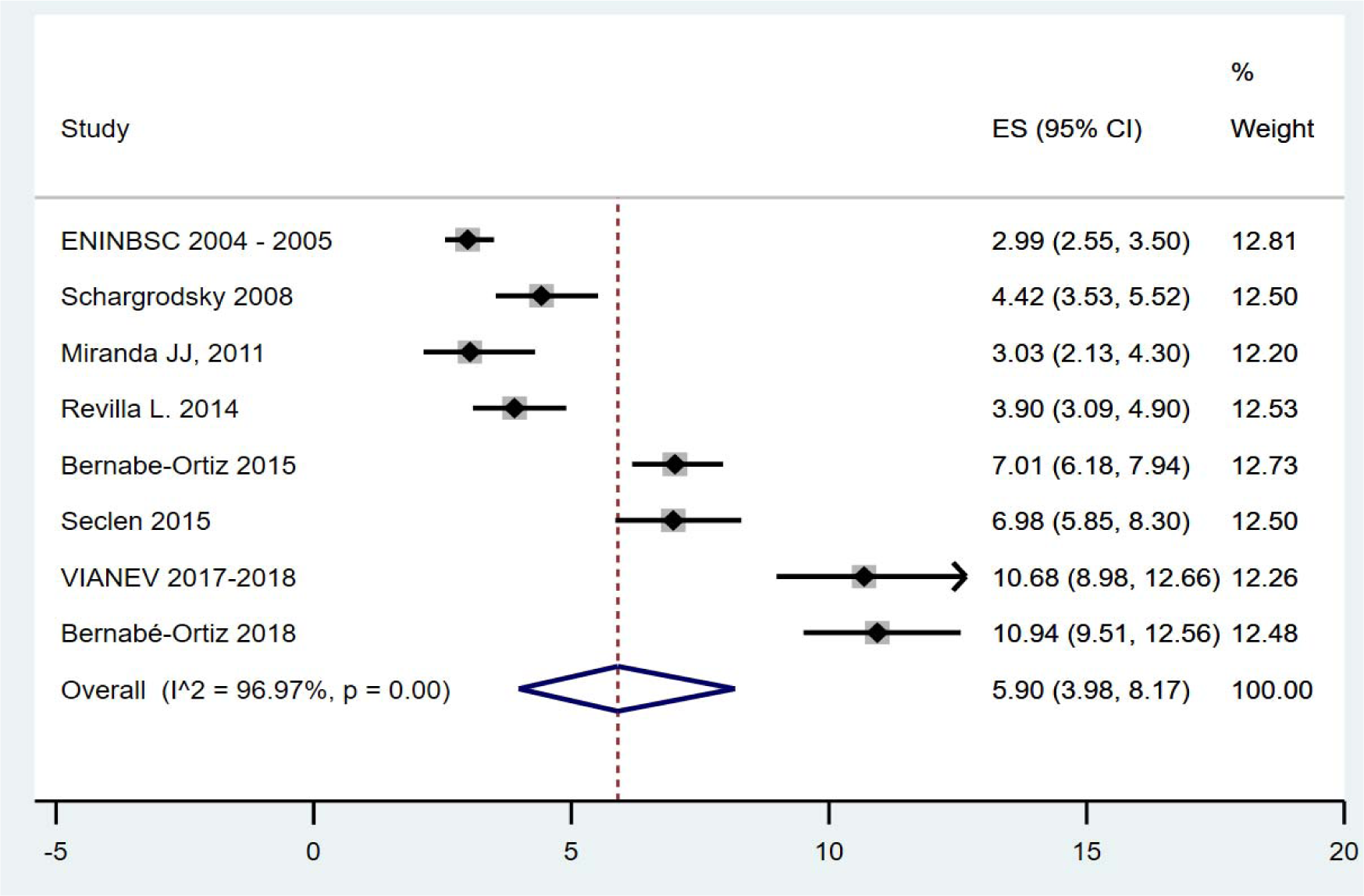
Pooled Prevalence of DM in Peru.

The sensitivity analysis of the prevalence of DM in Peru shows significant variations depending on the period and geographical scope of the included studies. Temporally, the prevalence of DM increased notably in studies conducted from 2013 onwards. Studies conducted before 2013 (n=3) reported an average DM prevalence of 3.43% (95% CI: 2.13 - 4.30) with considerable heterogeneity, indicated by an I² of 73.40%. In contrast, studies conducted from 2013 onwards (n=5) showed a significantly higher prevalence of 7.66% (95% CI: 5.36 - 10.32), with even higher heterogeneity (I² = 95.09%). Regarding geographical scope, subnational studies (n=6) presented a DM prevalence of 5.18% (95% CI: 3.94 - 8.02) with heterogeneity of I² = 95.22%, suggesting significant variations among the regions studied. On the other hand, national studies (n=2) reported a lower prevalence of 4.03% (95% CI: 3.54 - 4.54).

**Table 1.**
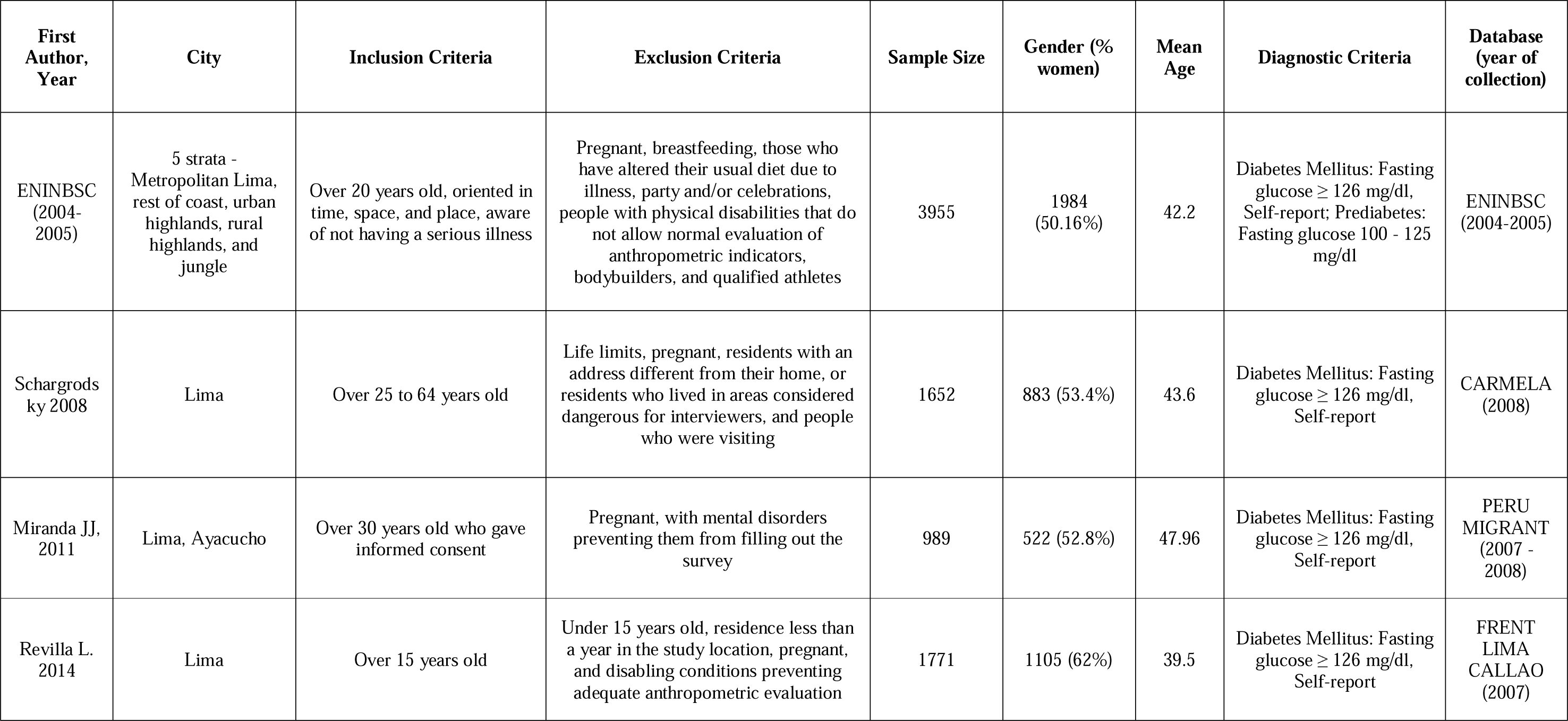
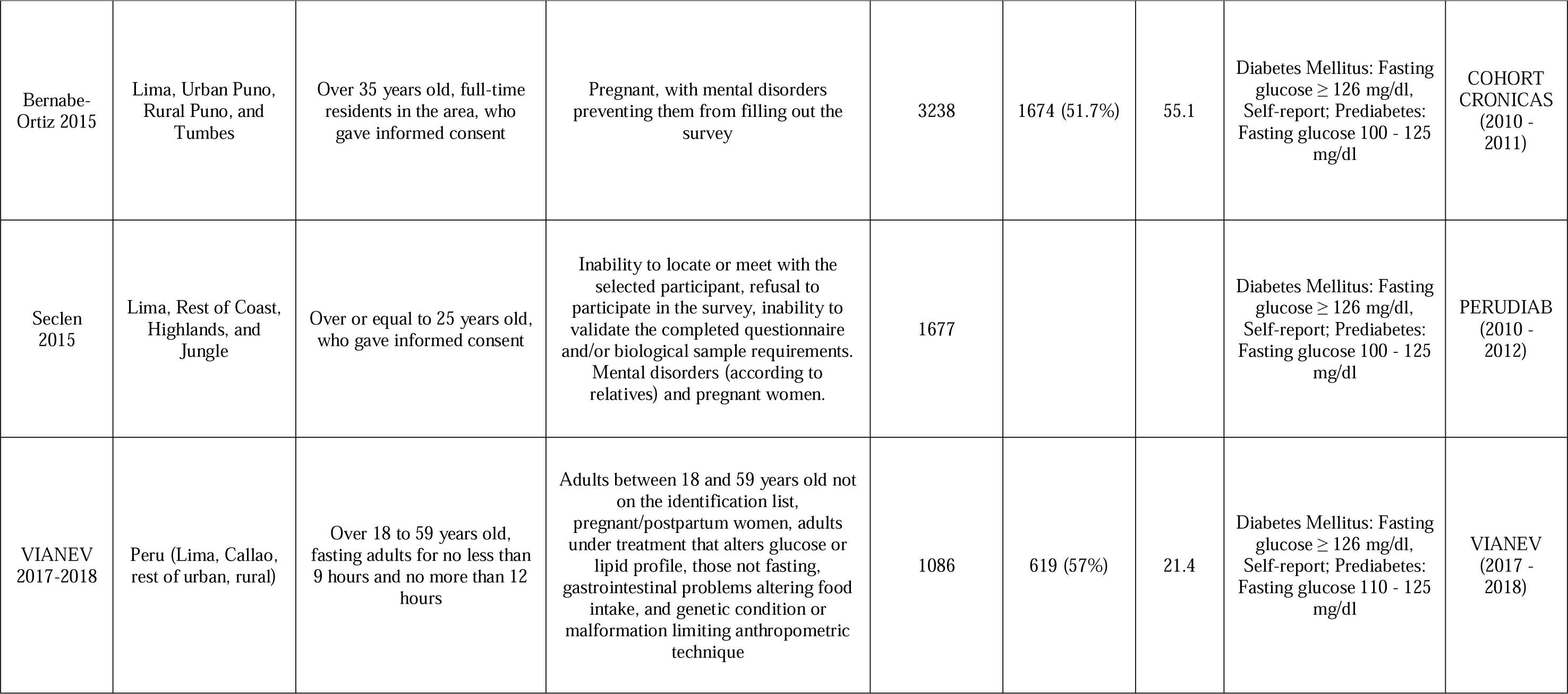
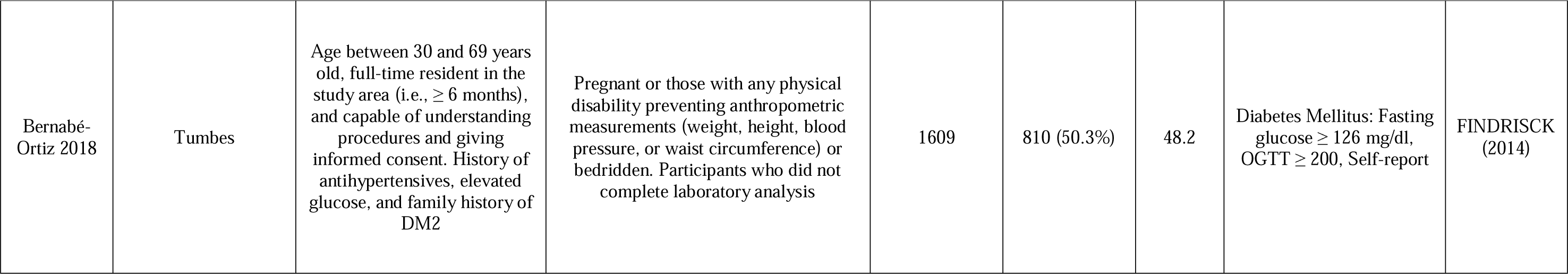
Summary of Studies on the Prevalence of Diabetes and Prediabetes in Peru.

**Table 2.**
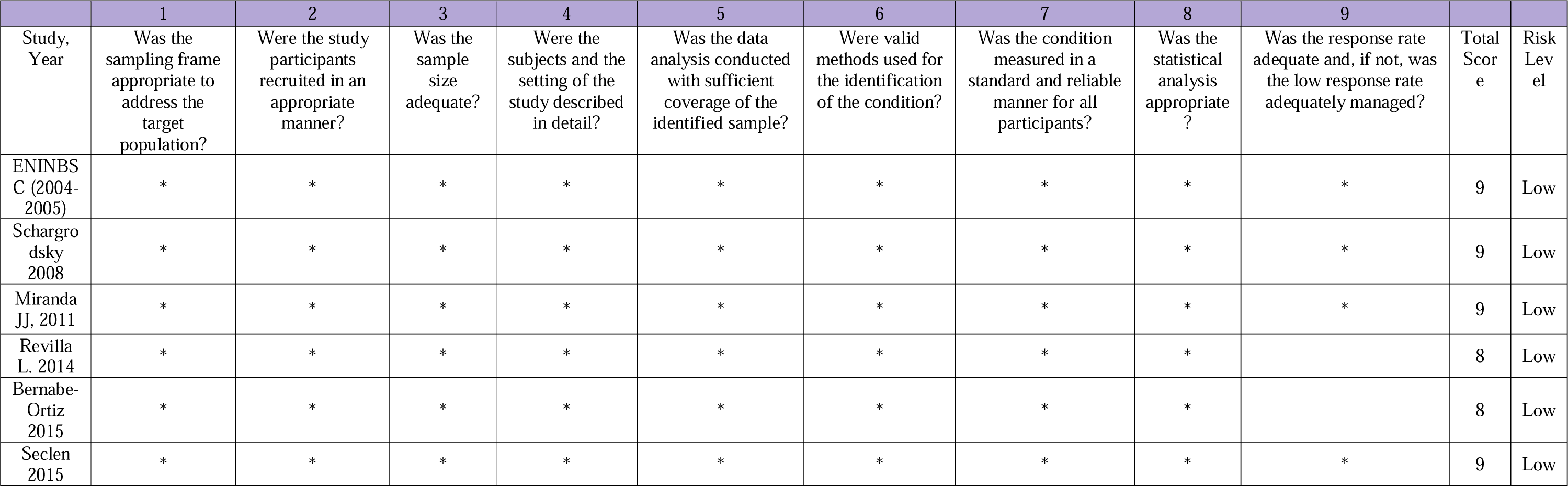
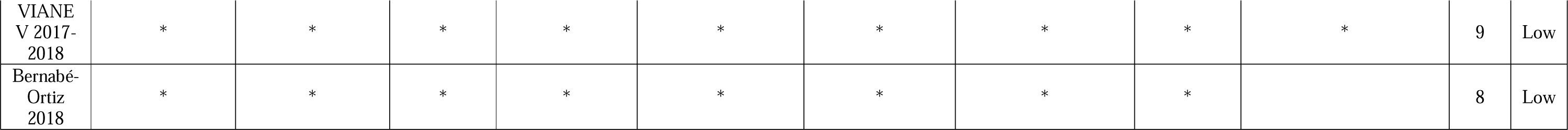
Risk of Bias in Studies on the Prevalence of Diabetes and Prediabetes in Peru.

**Table 3.** Sensitivity Analysis of DM prevalence in Peru.

|  | Number<br>of studies | Prevalence | IC 95% | I<sup>2</sup> |
| --- | --- | --- | --- | --- |
| **Time** |  |  |  |  |
| < 2013 | 3 | 3.43 | 2.13 - 4.30 | 73.40% |
| ≥ 2013 | 5 | 7.66 | 5.36 - 10.32 | 95.09% |
| **Scope** |  |  |  |  |
| Subnational | 6 | 5.18 | 3.94 - 8.02 | 95.22% |
| National | 2 | 4.03 | 3.54 - 4.54 |  |

The systematic review results indicate significant variations in the prevalence of DM, according to different categories. Ten studies were included for each sex, finding a prevalence of DM in men of 12.74% (95% CI: 5.93% - 23.22%, I² = 99.09%), while in women, the prevalence was considerably higher, at 42.9% (95% CI: 31.33% - 54.89%, I² = 99.23%). When analyzing the data by time, with two studies in each category, an increase in the prevalence of DM was observed in studies conducted from 2013 onwards (43.11%, 95% CI: 42.61% - 43.62%) compared to those conducted before 2013 (29.5%, 95% CI: 28.26% - 30.75%). Regarding geographical scope, the prevalence in subnational studies was 31.31% (95% CI: 29.59% - 33.05%), while national studies showed a slightly lower prevalence of 27.77% (95% CI: 27.33% - 28.21%). Table 4.

**Table 4.** Sensitivity Analysis of Prediabetes Prevalence in Peru.

|  | Number<br>of studies | Prevalence | IC 95% | I<sup>2</sup> |
| --- | --- | --- | --- | --- |
| **Time** |  |  |  |  |
| < 2013 | 2 | 4.32 | 3.76 - 4.92 |  |
| ≥ 2013 | 4 | 17.79 | 10.95 - 25.87 | 98.54% |
| **Scope** |  |  |  |  |
| Subnational | 4 | 13.54 | 7.29 - 21.35 | 98.68% |
| National | 2 | 6.49 | 5.81 - 7.21 |  |

### Meta-analysis of Prediabetes prevalence

The pooled prevalence for DM was 13.02% (95% CI: 6.31%—21.68%). However, heterogeneity was high (I² = 99.36%) (Figure 3).

**Figure 3.**
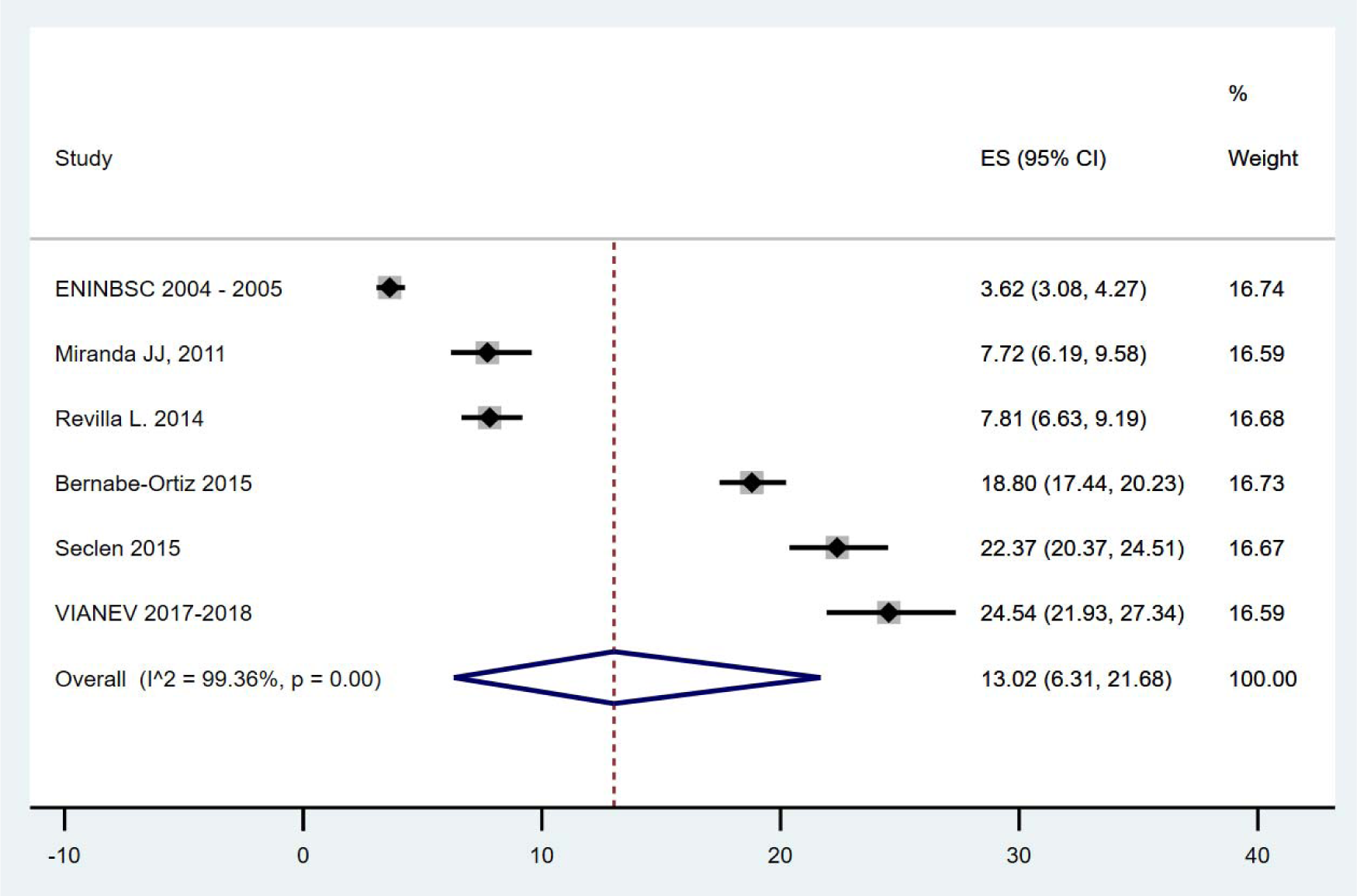
Pooled Prevalence of Prediabetes in Peru.

The sensitivity analysis of prediabetes prevalence in Peru also reveals significant differences. Temporally, there is an observed increase in the prevalence of prediabetes in more recent studies. For studies conducted before 2013 (n=3), the reported average prevalence was 3.43% (95% CI: 2.13 - 4.30), with a heterogeneity level of 73.40%. In comparison, studies conducted from 2013 onwards (n=5) showed a considerable increase in prevalence, reaching 7.66% (95% CI: 5.36 - 10.32), with a high degree of heterogeneity (I² = 95.09%). Regarding geographical scope, studies focusing on subnational regions (n=6) recorded a prediabetes prevalence of 5.18% (95% CI: 3.94 - 8.02), also with high heterogeneity (I² = 95.22%). National studies (n=2) showed a slightly lower prevalence of 4.03% (95% CI: 3.54 - 4.54), indicating less variability among results at the national level. Table 4.

## Discussion

### Prevalence of Diabetes and prediabetes in Peru

Our systematic review and meta-analysis revealed a prevalence of DM in Peru of 5.9%. These figures are consistent with those observed in other global studies, although they do not reach the extremely high levels reported in some regions ^(2,18)^. This relative moderation in prevalence suggests that, although DM and prediabetes are significant public health problems in Peru, the situation does not appear to be as severe as in countries with the highest prevalence rates.

However, prediabetes in Peru shows a prevalence of 13.02%, a concerning figure that suggests a substantial portion of the population is at risk of developing DM in the future. Comparatively, the global prevalence of prediabetes is also variable but is often reported in lower ranges than those found in this study ^(19)^. This finding underscores the urgent need for early intervention and prevention strategies in Peru to control the transition from prediabetes to diabetes.

It is crucial to recognize that although Peru does not reach the highest prevalence figures observed globally, the impact of DM and prediabetes on public health remains significant. Early detection, improvements in healthcare services, and awareness and education campaigns can be crucial in mitigating this problem before it reaches more alarming proportions.

### Studies with the Primary Objective of Understanding Diabetes/Prediabetes

Additionally, it is concerning to note the limited number of studies with the primary objective of explicitly determining the prevalence of DM and prediabetes in Peru. Of the studies analyzed, PERUDIAB ^(3)^ stood out as the only one exclusively focused on measuring these prevalences. Although other studies, such as VIANEV ^(9)^ and ENINBSC ^(7)^, included prevalence measurements, they did not focus solely on DM and prediabetes but covered a broader range of health indicators. This less targeted approach can influence the accuracy and focus of the reported prevalence measures.

Most other studies included in this review did not have the primary objective of studying DM or prediabetes. However, they leveraged glucose data collection conducted as part of their broader investigations to obtain these prevalence data. While this provides valuable information, it also raises questions about the consistency and prioritization of glucose measurement in these studies, which could affect the interpretation of the true disease burden in the population.

This situation underscores the need for more studies specifically designed to investigate the prevalence of DM and prediabetes in Peru. A more focused approach would improve data quality and allow for a more accurate assessment of trends and intervention needs, ensuring that public health policies are well-informed and effective in managing these chronic conditions.

### Inconsistency with DM Prevalence Findings

Another fundamental issue in our analysis is the inconsistency observed in results related to the prevalence of diabetes, which appears to have increased over time. For example, the study by Bernabé (2018), conducted in Tumbes (a region known for its high prevalence of this disease) ^(17)^, reported values as high as 10%. However, this result might not be representative of the entire country. Nevertheless, the VIANEV ^(9)^, a more recent national survey, also found a prevalence of 10% at the national level, suggesting an upward trend in the prevalence of diabetes throughout Peru.

On the other hand, the PERUDIAB ^(3)^ study reported a prevalence of 6.98%, which could be considered more in line with the national reality, although its scope was not entirely nationwide. This poses a significant dilemma: should we trust the results of VIANEV, a national survey, more, or the findings of PERUDIAB, which, while more specifically focused on diabetes, did not cover the entire country?

This dispersion of results complicates not only the understanding of the true prevalence of diabetes in Peru but also the planning and execution of effective public health interventions. The significant differences in reported prevalences could be attributable to various factors, such as methodological differences, real regional variations in disease prevalence, or even the possible evolution of diabetes over time.

### Inconsistency with Prediabetes Prevalence Findings

In addition to the previous case, prediabetes presents an even more concerning scenario due to the wide variations in reported values and the potential implications of these for public health. Results range from a low of 3.62% reported by ^(7)^ to a considerable 22.37% in the PERUDIAB ^(3)^ study and 24.54% in the VIANEV ^(9)^. However, the latter study presents a peculiarity that deserves special attention: the threshold used to define prediabetes was a fasting glucose level of 110 to 125 mg/dL.

If the VIANEV study had adopted the threshold recommended by the ADA ^(12)^, starting at 100 mg/dL, prevalence values could have escalated to approximately 40%, an extremely high figure potentially unrepresentative of the general situation in Peru. This adjustment in the cutoff point can dramatically alter the perception of the burden of prediabetes in the population, influencing the urgency and direction of intervention policies.

The disparity in these results underscores the importance of standardizing diagnostic methods and criteria used in epidemiological studies. Prediabetes, as a precursor state to diabetes, requires early monitoring and intervention to prevent progression to DM, a chronic condition with significant health and socioeconomic costs.

### Analysis of findings

This variability in the results of studies on DM and prediabetes in Peru prompts reflection on the concordance of the data obtained with estimates provided by global meta-analyses. While meta-analyses can offer a general overview of trends and prevalence, individual studies and national reports in Peru do not always reflect these findings. A plausible reason behind this discrepancy could be the diversity of methodological approaches and, in many cases, the lack of a primary focus exclusively on evaluating these conditions, except for the PERUDIAB study ^(3)^.

A significant gap in the literature is the absence of a clear, nationwide study specifically focused on the prevalence of diabetes and prediabetes. This raises serious concerns about our ability to effectively understand and address these chronic conditions in the Peruvian context. Without precise and representative data, it is challenging for policymakers and health professionals to design and implement effective interventions to mitigate the impact of diabetes and prediabetes on the population.

### Importance of the study for public health

Understanding diabetes and prediabetes is crucial for public healthcare. It provides essential data for planning healthcare policies. In Peru, where healthcare resources are limited, evaluating these chronic conditions helps policymakers prioritize resources more effectively. Further studies identify at-risk groups, which aids in targeting preventive measures to reduce disease rates and complications.

Health authorities need a clear picture of how widespread these diseases are so that they can design effective programs that address the needs of the affected populations. For example, public education campaigns promoting healthy lifestyles and early detection programs can be developed. Additionally, support systems for managing diabetes can be established. These strategies help prevent new cases and improve outcomes for those already living with diabetes, reducing the overall burden on the healthcare system and improving healthcare in society.

Well-designed studies on the prevalence of these conditions lay the groundwork for future research. They provide a basis for evaluating the effectiveness of current interventions and developing new evidence-based strategies. Moreover, these studies reveal trends over time and across different regions, which is essential for adapting health policies as demographics shift and health needs evolve. This research advances scientific knowledge and enhances a country’s readiness to address public health issues effectively. Therefore, such research is critically important for tackling public health challenges efficiently.

### Study Limitations

One of this study’s main limitations is the diversity of glucose measurement methods and diagnostic criteria used in the analyzed studies. This variability can significantly affect the comparability of results. While some studies use fasting glucose according to ADA criteria, others may use different thresholds, resulting in prevalences that are not directly comparable. This lack of uniformity complicates the interpretation of the true magnitude of diabetes and prediabetes at the national level.

Another important limitation is the uneven geographical coverage of the studies in the analysis. Many studies focus on urban areas or specific regions such as Tumbes or Lima and may not represent other regions of Peru, especially rural areas, or indigenous communities. This uneven distribution prevents the results from being generalizable to the entire Peruvian population, which could lead to an underestimation or overestimation of the national prevalence.

Most of the studies analyzed date back several years and may not reflect current trends in the prevalence of these metabolic conditions. The lack of more recent data prevents an analysis of the progress or worsening of the situation over time, which is crucial for the effective evaluation of health policies and programs implemented in recent years.

## Conclusions

The analysis of the prevalence of diabetes and prediabetes in Peru reveals a complex situation with significant variations in reported figures over time and across different regions. While the prevalence of diabetes appears to be increasing, the situation with prediabetes is particularly alarming, with estimates suggesting a large portion of the population is at imminent risk of developing diabetes. This study highlights the importance of addressing these chronic conditions with well-founded and targeted public health strategies.

To effectively address the rising prevalence of diabetes and prediabetes in Peru, it is imperative to adopt uniform and standardized diagnostic criteria in all future research. This standardization will enable valid comparisons between studies and a more accurate assessment of public health interventions. Additionally, it is recommended that national studies be conducted that include representations from all regions of the country, both urban and rural, to gain a comprehensive and detailed understanding of the situation. These studies should be complemented by continuous monitoring systems to track disease trends and the effectiveness of implemented policies.

Simultaneously, it is essential to prioritize developing and implementing early detection and prevention programs that promote healthy lifestyles and facilitate access to preventive health services. Such programs should be accompanied by educational and awareness campaigns highlighting the importance of regular glucose level checks, proper nutrition, and regular physical activity. These initiatives must be supported by sustained investment to ensure their long-term impact, aimed not only at reducing the prevalence of these conditions but also at improving the quality of life of the Peruvian population.

## Data Availability

All data produced in the present study are available upon reasonable request to the authors

## Acknowledgements

Special thanks to the Institute for Research in Tropical Diseases members, National University Toribio Rodríguez de Mendoza of Amazonas (UNTRM), Amazonas, Peru, for their support and contributions throughout this research.

## Informed Consent

Informed consent was not required for this study.

## Data Availability

Data are available upon request to the corresponding author.

## Author Contributions

### Víctor J. Vera-Ponce

Conceptualization, Investigation, Methodology, Resources, Writing - Original Draft, Writing - Review & Editing.

### Fiorella E. Zuzunaga-Montoya

Investigation, Project Administration, Writing - Original Draft, Writing - Review & Editing.

### Luisa Erika Vasquez-Romero

Investigation, Resources, Writing - Original Draft, Writing - Review & Editing.

### Joan A. Loayza-Castro

Software, Data Curation, Formal Analysis, Writing - Review & Editing.

### Enrique Vigil-Ventura

Validation, Visualization, Writing - Original Draft, Writing - Review & Editing.

### Willy Ramos

Methodology, Supervision, Funding Acquisition, Writing - Review & Editing.

---

**Supplementary file 1.**
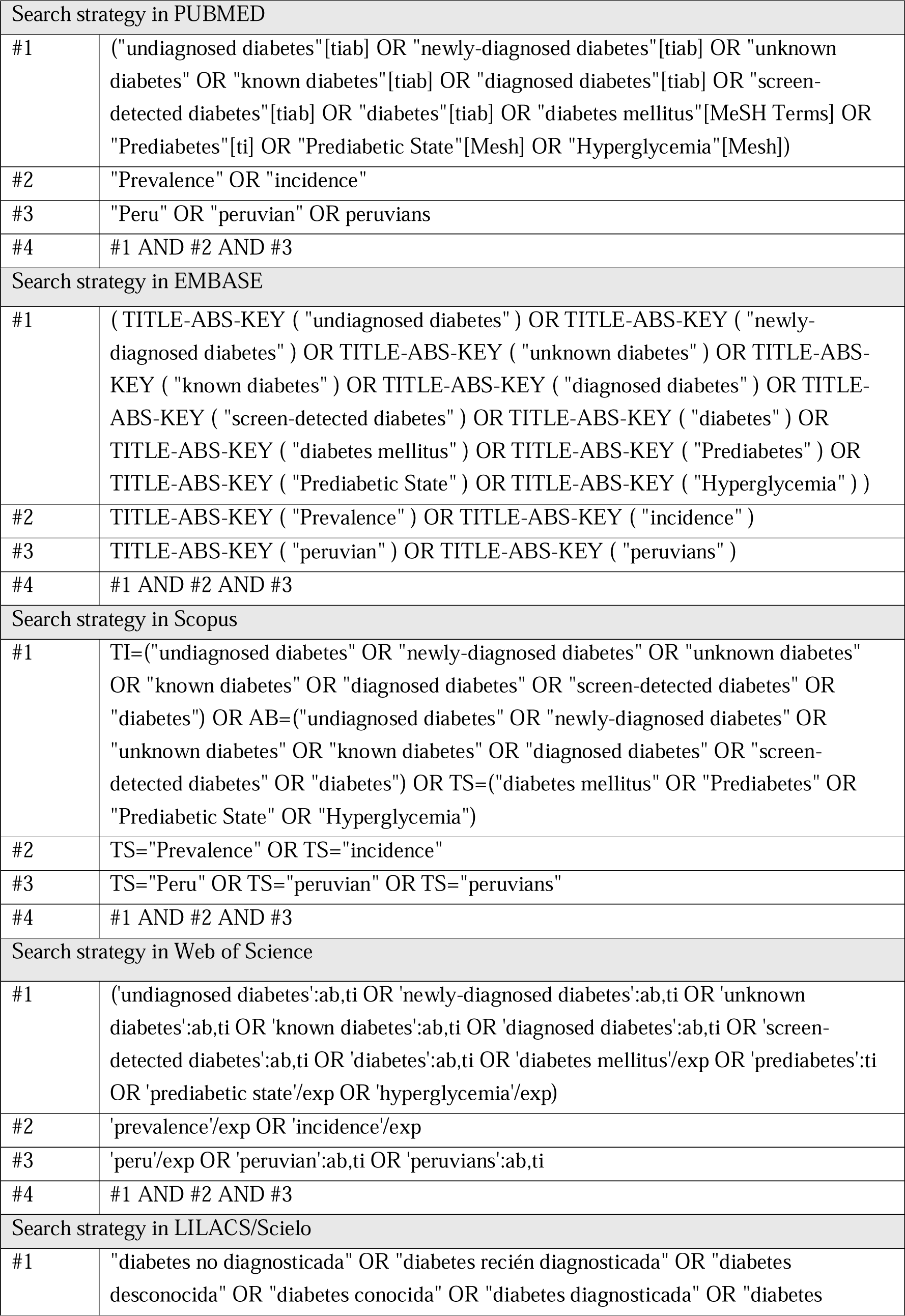
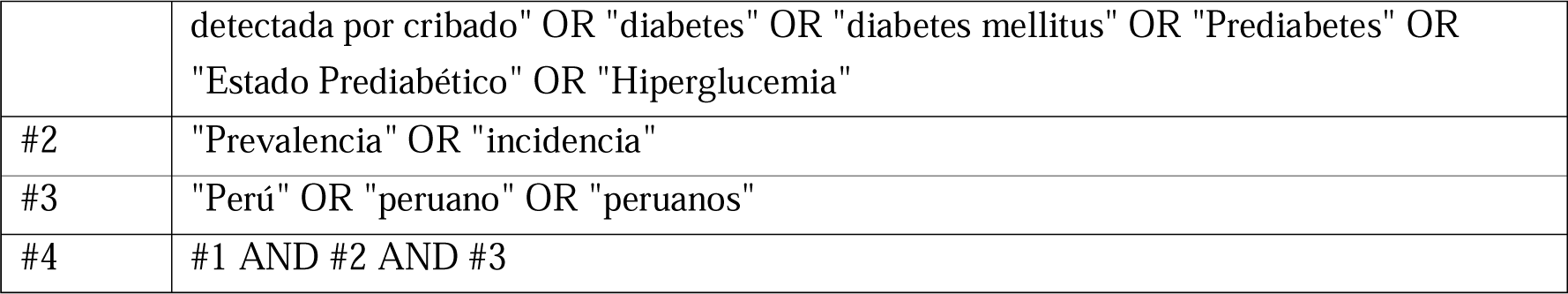

| Search strategy in PUBMED |  |
| --- | --- |
| #1 | ("undiagnosed diabetes"[tiab] OR "newly-diagnosed diabetes"[tiab] OR "unknown diabetes" OR "known diabetes"[tiab] OR "diagnosed diabetes"[tiab] OR "screen-detected diabetes"[tiab] OR "diabetes"[tiab] OR "diabetes mellitus"[MeSH Terms] OR "Prediabetes"[ti] OR "Prediabetic State"[Mesh] OR "Hyperglycemia"[Mesh]) |
| #2 | "Prevalence" OR "incidence" |
| #3 | "Peru" OR "peruvian" OR peruvians |
| #4 | #1 AND #2 AND #3 |
| Search strategy in EMBASE |  |
| #1 | ( TITLE-ABS-KEY ( "undiagnosed diabetes" ) OR TITLE-ABS-KEY ( "newly-diagnosed diabetes" ) OR TITLE-ABS-KEY ( "unknown diabetes" ) OR TITLE-ABS-KEY ( "known diabetes" ) OR TITLE-ABS-KEY ( "diagnosed diabetes" ) OR TITLE-ABS-KEY ( "screen-detected diabetes" ) OR TITLE-ABS-KEY ( "diabetes" ) OR TITLE-ABS-KEY ( "diabetes mellitus" ) OR TITLE-ABS-KEY ( "Prediabetes" ) OR TITLE-ABS-KEY ( "Prediabetic State" ) OR TITLE-ABS-KEY ( "Hyperglycemia" ) ) |
| #2 | TITLE-ABS-KEY ( "Prevalence" ) OR TITLE-ABS-KEY ( "incidence" ) |
| #3 | TITLE-ABS-KEY ( "peruvian" ) OR TITLE-ABS-KEY ( "peruvians" ) |
| #4 | #1 AND #2 AND #3 |
| Search strategy in Scopus |  |
| #1 | TI=("undiagnosed diabetes" OR "newly-diagnosed diabetes" OR "unknown diabetes" OR "known diabetes" OR "diagnosed diabetes" OR "screen-detected diabetes" OR "diabetes") OR AB=("undiagnosed diabetes" OR "newly-diagnosed diabetes" OR "unknown diabetes" OR "known diabetes" OR "diagnosed diabetes" OR "screen-detected diabetes" OR "diabetes") OR TS=("diabetes mellitus" OR "Prediabetes" OR "Prediabetic State" OR "Hyperglycemia") |
| #2 | TS="Prevalence" OR TS="incidence" |
| #3 | TS="Peru" OR TS="peruvian" OR TS="peruvians" |
| #4 | #1 AND #2 AND #3 |
| Search strategy in Web of Science |  |
| #1 | ('undiagnosed diabetes':ab,ti OR 'newly-diagnosed diabetes':ab,ti OR 'unknown diabetes':ab,ti OR 'known diabetes':ab,ti OR 'diagnosed diabetes':ab,ti OR 'screen-detected diabetes':ab,ti OR 'diabetes':ab,ti OR 'diabetes mellitus'/exp OR 'prediabetes':ti OR 'prediabetic state'/exp OR 'hyperglycemia'/exp) |
| #2 | 'prevalence'/exp OR 'incidence'/exp |
| #3 | 'peru'/exp OR 'peruvian':ab,ti OR 'peruvians':ab,ti |
| #4 | #1 AND #2 AND #3 |
| Search strategy in LILACS/SciELO |  |
| #1 | "diabetes no diagnosticada" OR "diabetes recién diagnosticada" OR "diabetes desconocida" OR "diabetes conocida" OR "diabetes diagnosticada" OR "diabetes |
|  | detectada por cribado" OR "diabetes" OR "diabetes mellitus" OR "Prediabetes" OR "Estado Prediabético" OR "Hiperglucemia" |
| #2 | "Prevalencia" OR "incidencia" |
| #3 | "Perú" OR "peruano" OR "peruanos" |
| #4 | #1 AND #2 AND #3 |

